# Do viral-associated pathways underlie the immune activation during the acute phase of severe major depression?

**DOI:** 10.1101/2024.11.05.24316765

**Authors:** Michael Maes, Yingqian Zhang, Kitiporn Plaimas, Apichat Suratanee, Jing Li, Abbas F. Almulla

## Abstract

**Background:** Major depressive disorder (MDD) and its most severe phenotype, major dysmood disorder (MDMD), are distinguished by the activation of the immune-inflammatory response system, T cell activation, and a relative T regulatory cell suppression. Nevertheless, these immune data were not used to characterize the features of the immune protein-protein interaction (PPI) network of MDMD.

**Objectives:** To identify the network’s nodes and bottlenecks as well as the biological processes that are overrepresented in the PPI network, we conducted PPI network, annotation, and enrichment analyses.

**Results:** The PPI network analysis has identified the following backbone genes: tumor necrosis factor-α (TNF), interleukin (IL)6, CXCL12, CXCL10, CCL5, cluster of differentiation (CD)4, CD8A, human leukocyte antigen (HLA)-DR, and FOXP3. A “cellular and defense response”, an “immune response system response”, and “a viral process that involves viral protein interaction with cytokines and cytokine receptors” were all highly associated with the network. The chemokine network and TNF and nuclear factor-κB (NFKB) pathways are additional biological pathways that are enriched in the PPI network. Molecular complex detection extracted one component from the data, including viral protein interaction with cytokine and cytokine receptors and “regulated by RELA” (an NFKB subunit).

**Conclusions:** Viral processes may underlie the activation of T cells and the cytokine and chemokine networks that are associated with MDMD. Future research on the pathogenesis of MDMD and MDD should examine whether and which viral infections are associated with the onset of these conditions, or whether viral reactivation is associated with the recurrence of illness.

## Introduction

Evidence now indicates that the acute phase of severe major depressive disorder (MDD) is a neuro-immune condition (Maes 1995, Almulla, Abbas Abo Algon et al. 2024). MDD consists of two primary subcategories: major dysmood disorder (MDMD) and simple dysmood disorder (SDMD) (Maes 2022, Maes, Moraes et al. 2023, Maes, Zhou et al. 2024). There exist clinical and biomarker-pathway distinctions between the two groups. MDMD, in contrast to SDMD, represents a more severe phenotype characterized by heightened levels of depression, anxiety, fatigue, and physiosomatic symptoms (Maes 2022, Maes, Moraes et al. 2023, Maes, Almulla et al. 2024). Moreover, a greater recurrence of illness, characterized by escalating episode frequencies and suicidal tendencies, distinguishes MDMD from SDMD. Nonetheless, a first episode of MDD can exhibit the MDMD phenotype if the severity of the illness is sufficient (Almulla, Abbas Abo Algon et al. 2024). Secondly, significant disparities exist in biomarkers and pathways between the two groups (Maes, Moraes et al. 2023). These disparities include elevated cytokine networks, enhanced T cell activation, diminished neurotrophic growth factors, heightened atherogenicity, and reduced antioxidant defenses (Maes, Rachayon et al. 2022, Maes, Rachayon et al. 2023, Maes, Zhou et al. 2024).

Investigations conducted on inpatients with severe MDMD revealed that this illness is marked by the activation of distinct cytokine profiles within the immune-inflammatory response system (IRS) and the compensatory immunoregulatory system (CIRS). The IRS indicates the activation of immunological and inflammatory networks, while the CIRS downregulates the IRS, thus averting an excessive IRS response and chronic inflammation, and facilitates a return to immune homeostasis or tolerance (Maes and Carvalho 2018). The MDMD inpatients exhibited markedly activated M1 macrophages, M2 macrophages, T helper (Th)-1 cells, Th-1 polarization, IRS, and CIRS profiles (Almulla, Abbas Abo Algon et al. 2024). The key cytokines, chemokines, and growth factors distinguishing MDMD from controls or predicting depression severity were interleukin (IL)-16, tumor necrosis factor (TNF)-related apoptosis-inducing ligand (TRAIL), IL-2 receptor (IL-2R), TNF-β, IL-6, and platelet-derived growth factor (PDGF). Furthermore, MDMD, as opposed to SDMD, is distinguished by particular alterations in immune cell CD markers, signifying T cell activation and the suppression of T regulatory (Treg) cells (Maes, Zhou et al. 2024). MDMD or its characteristics were markedly correlated with elevated numbers (or expression) of cells including CD4+ and CD8+, as well as CDL40+, CD71+, and HLA-DR+, in addition to cells expressing CD20+ and CB2 (cannabinoid receptors) (Maes, Rachayon et al. 2023). Conversely, there was a notable reduction in Treg cells, specifically CD4+CD25+FOXP3+CD152+, and CD4+CD25+FOXP3+GARP+ Treg cells (Maes, Zhou et al. 2024). These findings further suggest that MDMD is associated with an imbalance between IRS (T cell activation) and CIRS (Treg cell) activities (Maes and Carvalho 2018). It is important to emphasize that different phenotypes of depression, such as MDD in outpatients (Maes, Jirakran et al. 2024) or very mild SDMD (Maes, Vasupanrajit et al. 2023), exhibit significantly distinct immunological profiles.

However, no research has examined the molecular processes or pathways that underlie the alterations in IRS (M1, Th-1, Th-1 polarization), CIRS (M2), and alteration in immune cells in MDMD. This undertaking can be executed using protein-protein interaction (PPI) or gene-gene interaction analysis, followed by annotation and enrichment analysis (Maes, Plaimas et al. 2021, Gevezova, Sbirkov et al. 2023). Network analysis will reveal the hubs and bottleneck genes by calculating degree and betweenness centrality. This method may identify the most important drug targets for network modification and disorder treatment. Additionally, annotation and enrichment analysis will reveal potential underlying molecular mechanisms or intracellular networks that could function as novel therapeutic targets for the development of new treatments for the disorder (Maes, Plaimas et al. 2021).

Hence, this study was conducted to execute a network analysis of the PPI network of cytokines, chemokines, and growth factors identified in (Almulla, Abbas Abo Algon et al. 2024), alongside T cell markers from (Maes, Zhou et al. 2024). The network analysis will identify the hubs and bottlenecks of the shared network, while annotation and enrichment analysis will provide the ranking of the most significant molecular patterns of intracellular networks associated with immunological abnormalities in MDMD. Additionally, in the present investigation, we conducted functional enrichment analysis utilizing weighted scores based on the methodology established by Fan and Cui (2021).

Consequently, we utilized the mean expression values and the associated weights of all cytokines, chemokines, and growth factors quantified via a multiplex approach in the research conducted by (Almulla, Abbas Abo Algon et al. 2024). In addition, we conducted network and annotation studies on the differentially expressed cytokine, chemokine, and growth factor genes examined in (Almulla, Abbas Abo Algon et al. 2024), in conjunction with the differentially expressed CD markers identified in (Maes, Zhou et al. 2024).

### Participants

We enlisted 71 FE-MDMD patients from the psychiatric unit at Alhakeim Hospital in Najaf, Iraq, for a case-control study investigating cytokines, chemokines, and growth factors. Individuals with FE-MDMD exhibited an average disease duration of 2.5 months. Furthermore, the senior psychiatrist enlisted 40 healthy control participants who were acquaintances of patients, medical personnel, or associates of medical personnel from the same catchment area. The principal data are disseminated by (Almulla, Abbas Abo Algon et al. 2024). The flow cytometry study (Maes, Zhou et al. 2024) involved 30 outpatients with MDD (19 with MDMD and 11 with SDMD) at the psychiatric department of Chulalongkorn Hospital in Bangkok, Thailand, to examine T cell activation in MDMD. Posters and word of mouth were employed to recruit healthy volunteers from the same vicinity (Bangkok, Thailand) as the patients. This study incorporated 20 healthy controls.

Both studies employed identical diagnostic techniques and clinical inclusion and exclusion criteria. All participants were aged 18 to 65 years and received a DSM-5 diagnosis of MDD from a qualified psychiatrist, scoring a minimum of 17 on the Hamilton Depression Rating Scale (HAM-D) (Çakici, Sutterland et al. 2020). Almulla et al. (2024) exclusively recruited patients with MDMD (Almulla, Abbas Abo Algon et al. 2024). Maes et al. (2024) recruited individuals with MDD, categorized into MDMD and SDMD groups (Maes, Zhou et al. 2024). These diagnoses can be established via machine learning models developed from the Hamilton Depression Rating Scale (HAMD), the Hamilton Anxiety Rating Scale (HAMA), the recurrence of illness (ROI) index, or when the HAMD score is ≥ 22 and the HAMA score is ≥ 22. All patients included here were in the acute phase of MDMD, not in remission or partial remission.

The exclusion criteria were identical in both centers. We excluded participants who fulfilled the criteria for the following Axis I DSM-IV-TR disorders: psycho-organic disorders, dysthymia (unless part of double depression), bipolar disorder, schizoaffective disorder, schizophrenia, autism spectrum disorder, obsessive-compulsive disorder, generalized anxiety disorder, post-traumatic stress disorder, and substance use disorder (excluding nicotine dependence). The identical criteria were utilized for normal controls, who must also be devoid of a lifetime diagnosis of major depression and dysthymia, as well as a familial history of depression, mania, psychosis, substance use disorder, or suicide. Patients and controls with chronic liver or kidney disease, as well as pregnant and breastfeeding women, were excluded. Furthermore, we eliminated subjects with neurodegenerative, neuroinflammatory, or neurological disorders, including Alzheimer’s disease, Parkinson’s disease, stroke, and multiple sclerosis. Additionally, we excluded participants with (auto)immune diseases, including inflammatory bowel disease, rheumatoid arthritis, type 1 diabetes, cancer, psoriasis, scleroderma, systemic lupus erythematosus, chronic obstructive pulmonary disease, acute COVID-19 infection, severe or critical COVID-19 illness, long COVID, other infectious diseases such as influenza and hepatitis, asthma, cardiovascular disorders, and a history of allergic or inflammatory responses within the preceding three months. Additionally, we excluded participants who received immunosuppressive or immunomodulatory therapies, including glucocorticoids, those who had recently (within the last three months) consumed nonsteroidal anti-inflammatory drugs (NSAIDs) or other analgesics, and individuals on therapeutic doses of antioxidants or omega-3 supplements.

### Ethics statement

This work constitutes a secondary data analysis utilizing open, deidentified, and non-coded data sets; hence, it qualifies as non-human subjects research exempt from IRB approval. In our prior case-control research (Almulla, Abbas Abo Algon et al. 2024, Maes, Zhou et al. 2024), we detected significantly differentially expressed proteins (DEPs) in peripheral blood immune cells and in cytokines/chemokines/growth factors/CD markers compared to normal controls. Both case-control studies obtained permission from our local Institutional Review Boards, and all participants provided written informed consent prior to their involvement in the study. The Institutional Review Board of the College of Medical Technology of the Islamic University of Najaf, Iraq, sanctioned the study under Document No. 18/2021. The study protocol of Maes et al. (2024) received approval from the Institutional Review Board of Chulalongkorn University, Faculty of Medicine, Bangkok, Thailand (#528/63). The study was executed in accordance with Iraqi, Thai, and international ethical and privacy standards, including, but not limited to, the World Medical Association’s Declaration of Helsinki, the Belmont Report, the CIOMS Guidelines, and the International Conference on Harmonization of Good Clinical Practice. Furthermore, our institutional review board rigorously complies with the International Council for Harmonisation Good Clinical Practice (ICH-GCP) guidelines for human research safety.

### Biochemical assays

In both investigations, blood samples were collected between 8:00 and 11:00 a.m. with a disposable syringe and serum tubes. Following centrifugation of the blood at 3500 rpm, serum was isolated and stored in Eppendorf tubes at -80 °C in tiny aliquots until thawed for biomarker testing. The interested reader is directed at our publications detailing the analytical methodologies (Almulla, Abbas Abo Algon et al. 2024, Maes, Zhou et al. 2024).

WEAT analysis.

*Weat analysis performed using cytokine/chemokine/growth factors in MDMD*.

Using the method of (Fan and Cui 2021) (https://www.cuilab.cn/weat/), we performed functional enrichment analysis using weighted scores obtained from all variables in the study by (Almulla, Abbas Abo Algon et al. 2024). The gene identification of the cytokines/chemokines/growth factors that were analyzed in the initial phase of the study is depicted in **Table 1**. This table also displays the average expression values of the measurements in MDMD patients and controls. Using the formula average_MDMD – average_controls, we determined the weight (or essentiality score) of each gene based on these discrepancies, with higher scores indicating greater essentiality or importance for MDMD. As a result, we employed the weights specified in Table 1 to conduct Weighted Enrichment Analysis Tools (WEAT) (available at https://www.cuilab.cn/weat/). Our results section (refer to the below) will display the results of the Kyoto Encyclopedia of Genes and Genomes (KEGG) pathway enrichment analyses and Gene Ontology (GO) enrichment (https://genome.jp/kegg/).

**Table 1.** Average (Avg) expression of the serum cytokines, chemokines and growth factors measured in patients with major dysmood disorder (MDMD) and healthy controls (HC) and their computed weights, which are used in enrichment analysis.

| Name | Gene ID | Avg in HC | Avg in MDMD | Weight |
| --- | --- | --- | --- | --- |
| C-C motif chemokine ligand 27 | CCL27 | -0.993200333 | 0.387590374 | 1.380791 |
| C-C motif chemokine ligand 11 | CCL11 | -0.498178032 | 0.19441094 | 0.692589 |
| Colony stimulating factor 3 | CSF3 | 0.051415714 | -0.020064669 | -0.07148 |
| Colony stimulating factor 2 | CSF2 | -0.340418184 | 0.132846121 | 0.473264 |
| C-X-C motif ligand 1 | CXCL1 | -0.522932743 | 0.204071314 | 0.727004 |
| Hepatocyte growth factor | HGF | -0.968750474 | 0.378048965 | 1.346799 |
| Interferon gamma | IFNG | -0.222619502 | 0.086875903 | 0.309495 |
| Interleukin 1-alpha | IL1A | 0.081109168 | -0.031652358 | -0.11276 |
| Interleukin 1-beta | IL1B | 0.166606711 | -0.065017253 | -0.23162 |
| Interleukin 1 receptor antagonist | IL1R1 | -0.670766609 | 0.261762579 | 0.932529 |
| Interleukin 2 | IL2 | 0.065999419 | -0.025755871 | -0.09176 |
| Interleukin 2 receptor | IL2RA | -0.827629261 | 0.322977273 | 1.150607 |
| Interleukin 4 | IL4 | -0.276729039 | 0.10799182 | 0.384721 |
| Interleukin 5 | IL5 | 0.212457109 | -0.082910091 | -0.29537 |
| Interleukin 6 | IL6 | -0.505301334 | 0.197190765 | 0.702492 |
| C-X-C motif ligand 8 | CXCL8 | -0.108893613 | 0.042495068 | 0.151389 |
| Interleukin 9 | IL9 | -0.702492752 | 0.274143513 | 0.976636 |
| Interleukin 10 | IL10 | -0.059922232 | 0.023384286 | 0.083307 |
| Interleukin 12 | IL12P70 | -0.260999911 | 0.101853624 | 0.362854 |
| Interleukin 13 | IL13 | -0.26312619 | 0.102683391 | 0.36581 |
| Interleukin 15 | IL15 | -0.275978897 | 0.107699082 | 0.383678 |
| Interleukin 16 | IL16 | -1.17127045 | 0.457081151 | 1.628352 |
| Interleukin 17 | IL17A | -0.291181273 | 0.113631716 | 0.404813 |
| Interleukin 18 | IL18 | -0.679701455 | 0.265249348 | 0.944951 |
| C-X-C motif ligand 10 | IP10 | -0.548759209 | 0.214149935 | 0.762909 |
| Leukemia inhibitory factor | LIF | -0.2147785 | 0.083816 | 0.298595 |
| C-C Motif Chemokine Ligand 2 | CCL2 | -0.520846587 | 0.203257205 | 0.724104 |
| C-C Motif Chemokine Ligand 7 | CCL7 | -0.246266484 | 0.096103994 | 0.34237 |
| Colony stimulating factor 1 | CSF1 | -0.992314209 | 0.387244569 | 1.379559 |
| Macrophage migration inhibitory factor | MIF | -0.191819097 | 0.074856233 | 0.266675 |
| C-X-C motif ligand 9 | CXCL9 | -0.430871698 | 0.168145053 | 0.599017 |
| C-C Motif Chemokine Ligand 3 | CCL3 | 0.192464412 | -0.075108063 | -0.26757 |
| C-C Motif Chemokine Ligand 4 | CCL4 | -0.711276431 | 0.27757129 | 0.988848 |
| Nerve Growth Factor-beta | BNGF | -0.394633987 | 0.154003507 | 0.548637 |
| Platelet-derived growth factor | PDGFBB | -0.999472898 | 0.390038204 | 1.389511 |
| C-C Motif Chemokine Ligand 5 | CCL5 | -0.840278477 | 0.327913552 | 1.168192 |
| Stem cell factor | KITLG | -0.679492209 | 0.265167691 | 0.94466 |
| Stem cell growth factor | SCGFB | -1.1196397 | 0.436932566 | 1.556572 |
| C-X-C motif ligand 12 | CXCL12 | -0.735480778 | 0.287016889 | 1.022498 |
| Tumor necrosis factor-alpha | TNF | -0.506191239 | 0.197538045 | 0.703729 |
| Lymphotoxin alpha | LTA | -0.721703817 | 0.281640514 | 1.003344 |
| TNF Superfamily Member 10 | TNFSF10 | -1.04690335 | 0.408547649 | 1.455451 |
| Vascular endothelial growth factor | VFGFA | 0.144462726 | -0.056375698 | -0.20084 |

### Selection of seed proteins in our network analysis

STRING 11.9 (https://string-db.org) and Cytoscape (https://cytoscape.org) modules, including NetworkAnalyzer (Cytoscape App Store - NetworkAnalyzer), were employed to conduct the network analysis. The minimum required interaction score was 0.400, and the set organism was homo sapiens. We generated PPI networks with zero order, which consisted solely of the seed proteins. The latter was composed of the DEPs with respect to cytokines/chemokines/growth factors and CD markers as well. The DEPs utilized in this analysis are listed in Table 1. As a result, we analyzed the network characteristics of the zero network, which included the number of nodes and edges, network diameter and radius, clustering coefficient, and network density. Hub nodes were authenticated as the top nodes with the highest degree value, while top non-hub bottlenecks were identified as nodes with the highest betweenness centrality. The network’s backbone is formed by both the DEPs with the highest degree and the non-hub bottlenecks.

### Annotation and Enrichment analysis

In addition to WEAT analysis performed on the weights of all cytokines/chemokines/growth factors, we also analyzed the DEPs from the combined (Almulla, Abbas Abo Algon et al. 2024, Maes, Zhou et al. 2024). The combined DEPs, which include cytokines/chemokines/growth factors/CD markers, were subjected to annotation-enrichment analyses. Therefore, in those analyses, we integrate the significant M1, Th1, Th2, IRS, and CIRS cytokines/chemokines and growth factors with CD markers of T cell activation, B cells bearing CB receptors, and T reg cells. We used False Discovery Rate (FDR) p correction to adjust for multiple associations.

The top 100 most relevant genes were included in an extended network, and the DEPs were applied to Enrichr (https://maayanlab.cloud/Enrichr/). The DEPs were searched against KEGG 2021 Human pathways, and the Transcriptional Regulatory Relationships Unraveled by Sentence-based Text mining (TRRUST) factors. The results were presented in the form of bar graphs that were generated using Appyters to display the top 10 performing enriched terms. In these bar graphs, the statistical significance is denoted by the asterisks located adjacent to the p-values.

Furthermore, the DEPs network was subjected to enrichment analysis of KEGG pathways and disease-gene associations using STRING. The latter method included the top 10 processes that were displayed using functional enrichment visualization sorted by -log (FDR) and grouped by >0.8 similarity.

The list of seed genes was supplemented with known protein interactions from Metascape (http://metascape.org). The combined GO molecular processes, immunologic signatures, and KEGG and Reactome (https://reactome.org) pathways were used to conduct enrichment analysis (min network size = 3, maximal network size = 5, beta testing, min enrichment = 1.5, set species = homo sapiens). Based on Kappa-statistical similarities among their gene memberships, Metascape automatically clusters the significant terms into a tree in a hierarchical fashion. This is advantageous due to the substantial overlap between the terms employed, which may otherwise result in a significant amount of redundancy in the output terms. The molecular complex detection (MCODE) method was employed as the primary analysis to identify small components of densely connected nodes, which are indicative of molecular complexes (Bader and Hogue 2003). Biological meanings were extracted from the network components and GO enrichment was applied. The top 3 best p-value terms are listed.

Using the module explorer in OmicsNet (https://www.omicsnet.ca), we constructed a Network with MultiOmics Enrichment Analysis. The inventory of seed DEPs was expanded to include known protein interactions from STRING. The OmicsNet function regulation explorer was employed to establish Panther biological processes (www.pantherdb.org) and KEGG gene-protein processes. The module explorer was used to delineate modules in the enlarged network. We created a 3-dimensional network graph that also displays the network’s backbone.

## Results

### The PPI network topography of FE-MDMD

To represent the protein interactions in FE-MDMD, an undirected network was constructed by utilizing all seed genes (cytokine/chemokine/growth factor/CD marker DEPs). The zero-order network was composed of 34 nodes, as illustrated in Table 1. However, there were three singletons that were not part of the PPI network that had been established. Subsequently, these were eliminated: CNR1, CNR2, and CNGB1. The zero-order protein network, depicted in **Figure 1**, contained 31 nodes. The number of edges (n=348) exceeded the anticipated number of edges (n=55) with a p-enrichment value of 1.0e-16. The average local clustering coefficient was 0.86, and the average node degree was 22.5. The characteristic path length was 1.252, the network diameter was 2, the network radius was 1, and the network density was 0.748. The main hubs were TNF (degree=31), IL-6 (29), CD4 (29), CD8A (28), CCL5 (28), CXCL10 (28), and CXCL12 (27) according to the network degree analysis. No other non-hub bottlenecks were included in the top 5 bottleneck nodes. Consequently, TNF, IL-6, CD4, CD8A, CCL5, CXCL10, and CXCL12 constitute the zero-order network’s backbone.

**Figure 1.**
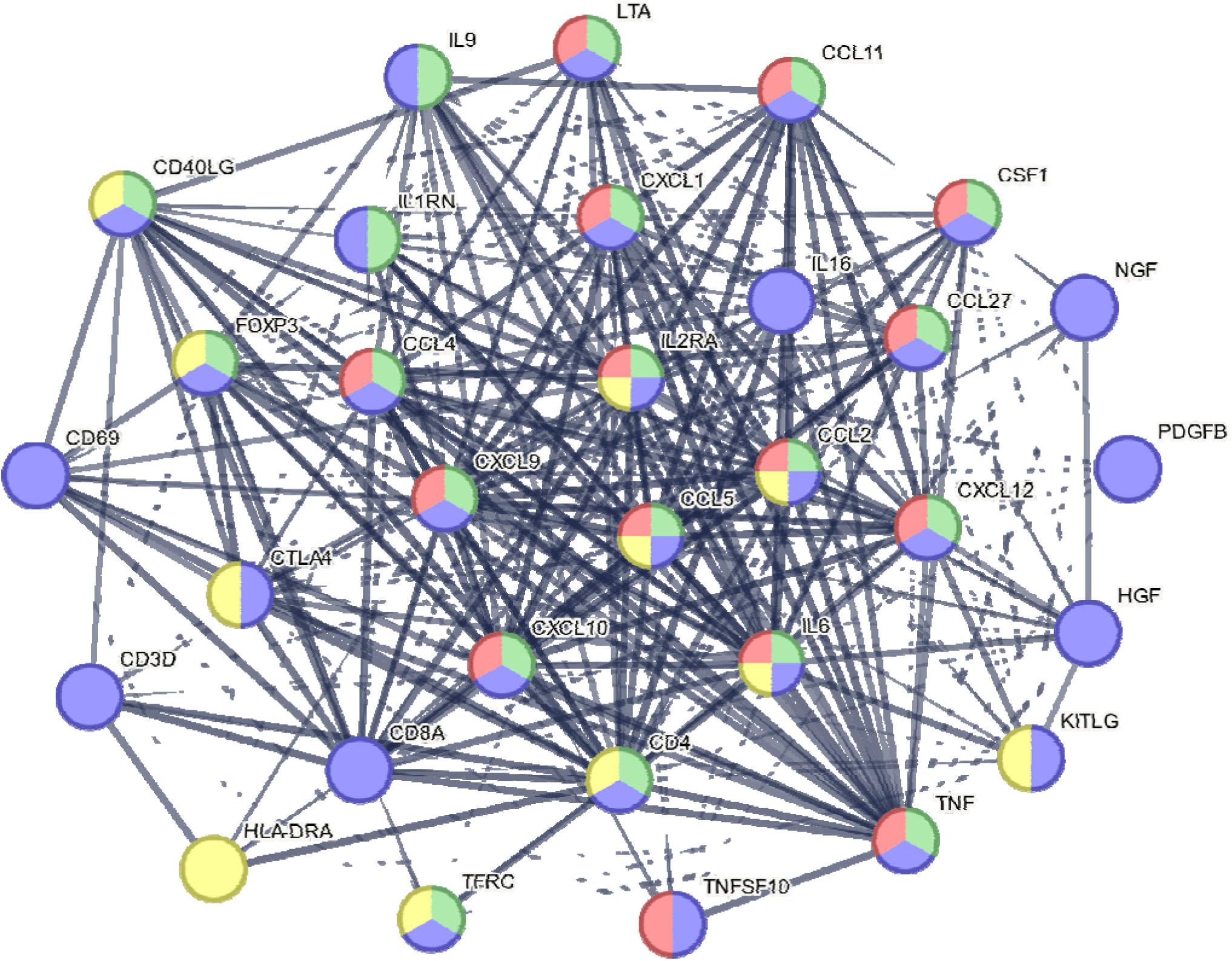
Zero-order protein-protein interaction network of the differently expressed cytokine / chemokine/ growth factor/CD marker genes of major dysmood disorder. Yellow: regulation of T cell activation, Red: Viral protein interaction with cytokine an cytokine receptor, Green: Defense response, Pink: cellular response to a stimulus, Al nodes: response to a stimulus.

Consequently, the list of 31 seed genes was expanded (first shell only) to include known protein interactions from STRING. The extended network consists of 217 nodes and 5907 edges, whereas the anticipated number of edges was 1428 (PPI enrichment p value of 1.0 e-16). The average local clustering coefficient was 0.678, and the average node degree was 54.4. The module explorer of OmicsNET revealed the presence of 5 significant modules. The first module (n=54, 1.81e-17) was centered around chemokines, particularly CXCL12 and CCL5, the second module (n=42, 9.28e-18) was centered around TNF, the third module (n=22, 1.25e-0.5) was centered around CD4, the fourth module centered around HLA-DR (n=21, p=3.58e-0.5), and the fifth module was centered around FOXP3 (n=14, p=0.00463). The degree analysis indicated the top hubs as TNF (53), CD4 (47), CXCL12 (38), CCL5 (37), CCL11 (25) and HLA-DR (22). FOXP3 was the third most significant bottleneck, following CD4 and TNF, and preceding CCL5 and CXCL12.

### Weat enrichment

The results of the WEAT enrichment analysis conducted on the weights of all cytokines/chemokines/growth factors obtained in the (Almulla, Abbas Abo Algon et al. 2024) study is presented in **Table 2**. We conducted a search of the network against the KEGG pathways. The cytokine-cytokine receptor interactions, viral protein interaction with cytokine and cytokine receptor, TNF signaling pathway, and chemokine signaling pathway were the top pathways (FDR all < e-9).

**Table 2.**
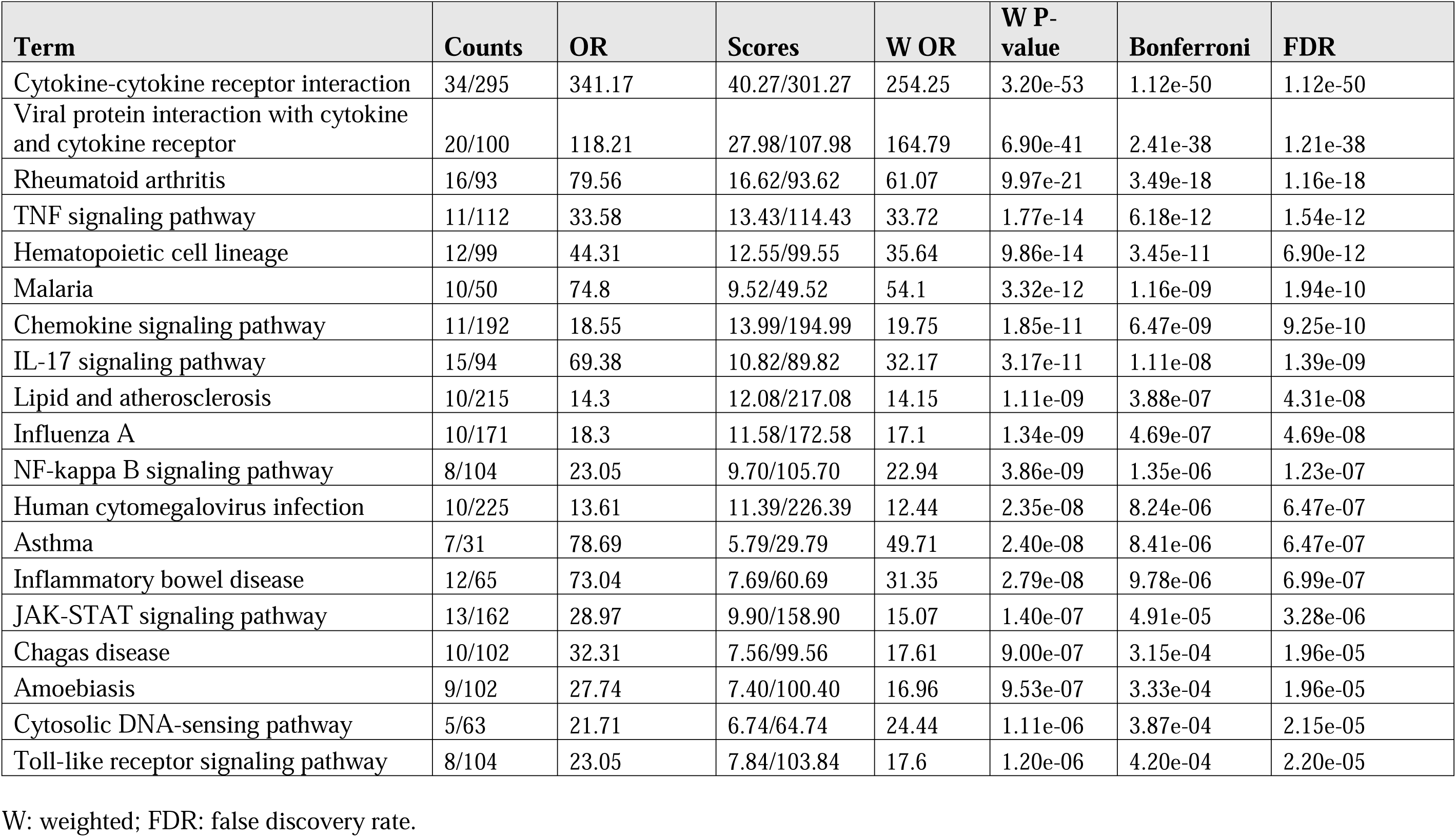
Results of Weighted Enrichment Analysis Tools (WEAT) with the KEGG pathways associated with major dysmood disorder.

### STRING enrichment

Additionally, we conducted a search of the zero-order network using the 31 DEPs shown in Figure 1 against the KEGG pathways and disease-gene associations using STRING. The results of the KEGG enrichment are illustrated in **Figure 2**. The KEGG pathways that were most enriched in the zero-network were the viral protein interaction with cytokine and cytokine receptor, as well as the cytokine-cytokine receptor interactions. The chemokine signaling pathway followed at a distance. The nuclear factor-κB and TNF signaling pathways were the fourth and fifth most significant pathways.

**Figure 2.**
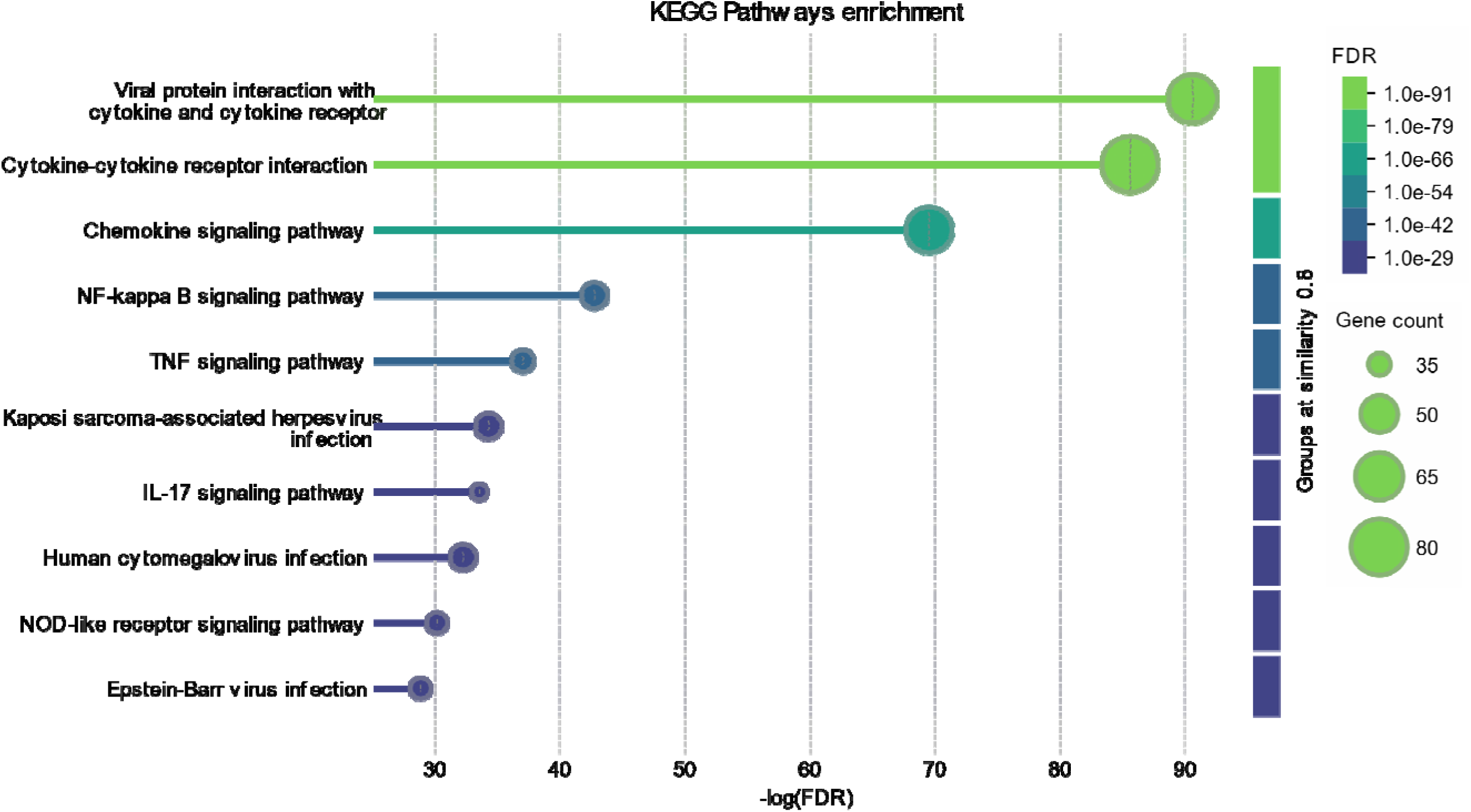
Functional enrichment visualization using Kyoto Encyclopedia of Genes and Genomes (KEGG) pathway. Used are the differently expressed cytokine / chemokine/ growth factor/ CD marker genes of major dysmood disorder.

In Figure 1, we have projected a selection of GO functions, including the response to a stimulus (31/6357, 2.58e-10), regulation of T cell activation (11/376, 2.63e-9), defense response (20/1394, 5.35e-13), cellular response to a stimulus (30/6357, 2.58e-11), and one KEGG pathway, namely viral protein interaction with cytokine and cytokine receptor (15/96, 3.14e-24). The enrichment of disease-gene associations is illustrated in **Figure 3**. This analysis suggests that the zero-order network is substantially enriched in autoimmune disease, primary immunodeficiency disease, and immune system disease. A variety of viral infection diseases are included, in addition to inflammatory disorders.

**Figure 3.**
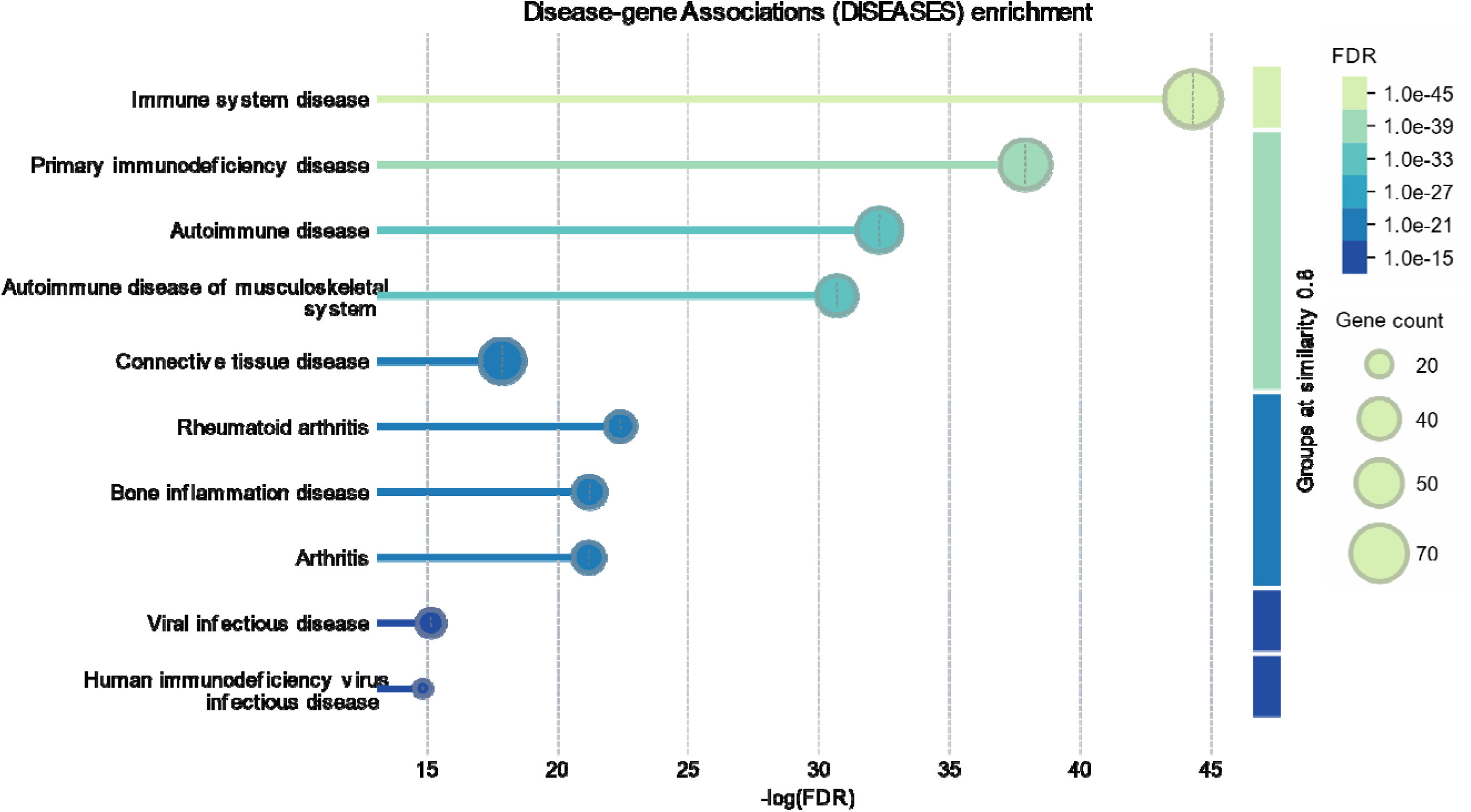
Functional enrichment visualization of Disease-Gene Associations. Used are the differently expressed cytokine / chemokine/ growth factor/ CD marker genes of major dysmood disorder.

### Enrichr enrichment

The results of the Enrichr enrichment analysis (extended networks) search against KEGG pathways and TTRUST transcription factors are depicted in **Figure 4** and **Figure 5**, respectively, as bar graphs. The KEGG pathway analysis yielded comparable results to the KEGG analysis conducted on the zero-order network and the WEAT analysis and additionally indicated that the Toll-Like Receptor (TLR) pathway was significantly enriched in the network. NFKB1 (nuclear factor (NF)-κB), REL (REL Proto-Oncogene, NF-κB Subunit), and RELA were the most important transcriptional factors associated with the Enrichr extended network (Figure 5).

**Figure 5.**
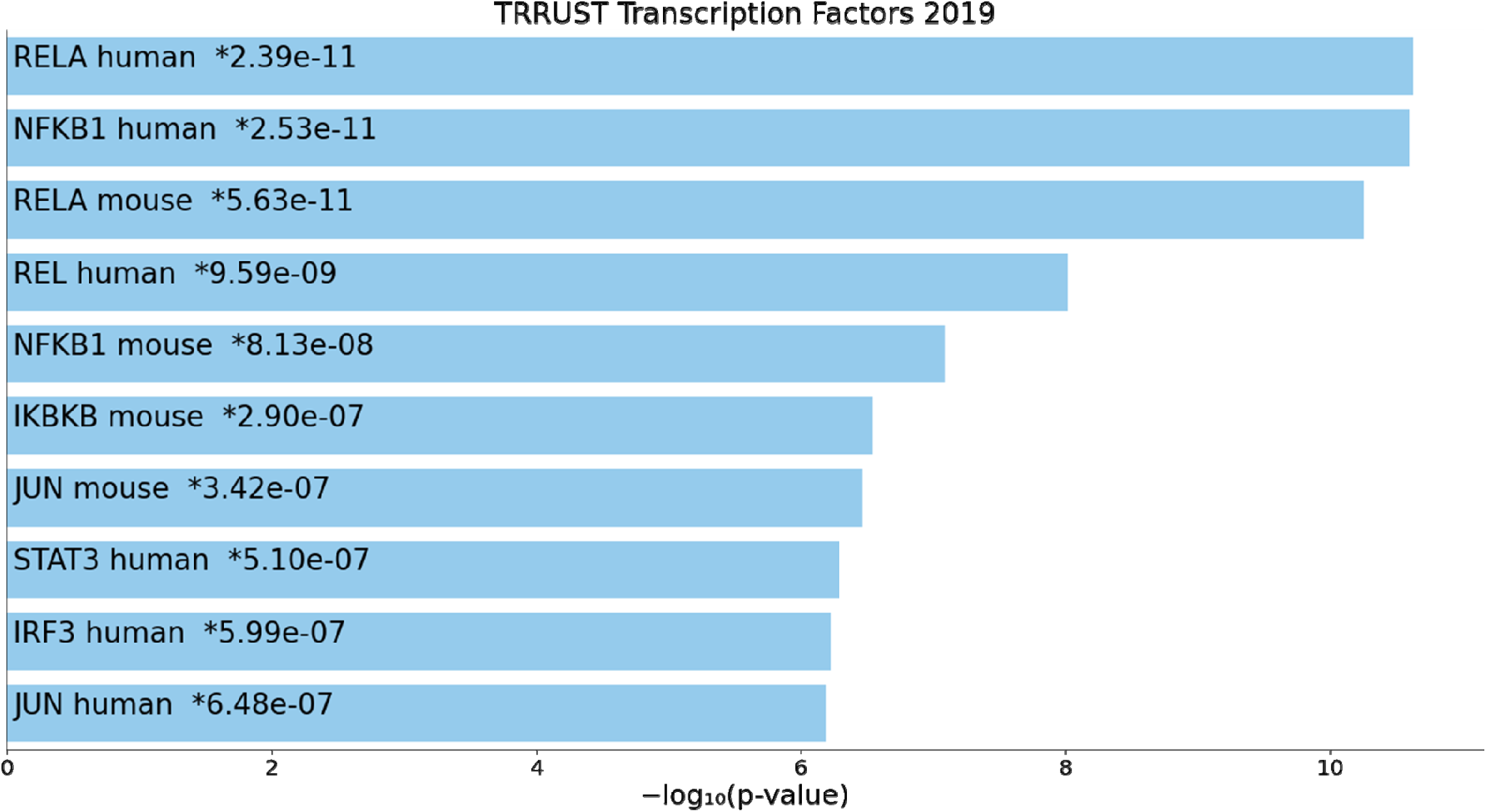
Heatmap (top-10) of the Transcriptional Regulatory Relationships Unraveled by Sentence-based Text mining (TRRUST) transcription factors accumulated in the differently expressed genes of the zero-order network of major dysmood disorder.

### Metascape enrichment

MCODE analysis performed using an extended network constructed with Metascape showed that one molecular complex could be extracted from the data, namely cytokine-cytokine receptor interaction, viral protein interaction with cytokine and cytokine receptor, and regulated by RELA (see **Table 3**).

**Table 3.** Results of Molecular Complex Detection (MCODE) analysis performed on the differently expressed genes of major dysmood disorder.

| Networks | Pathway | GO | Annotation (Log10P) |
| --- | --- | --- | --- |
| MCODE_All | KEGG | hsa04060 | Cytokine-cytokine receptor interaction (-34.9) |
|  | KEGG | hsa04061 | Viral protein interaction with cytokine and cytokine receptor (-29.2) |
|  | TRRUST | TRR01158 | Regulated by RELA (-21.1) |
| SUB MCODE_1 | KEGG | hsa04060 | Cytokine-cytokine receptor interaction (-34.9) |
|  | KEGG | hsa04061 | Viral protein interaction with cytokine and cytokine receptor (-29.2) |
|  | TTRUST | TRR01158 | Regulated by RELA (-21.1) |

### OmicsNET enrichment

Figure 6 illustrates the enrichment visualization of the extended network based on the critical cytokine/chemokine/growth factor/CD marker DEPs, as described in the “The PPI network topography of FE-MDMD” section. Using a perspective architecture, this figure illustrates the nodes of the five significant modules that were constructed using the OmicsNET module explorer. The backbone nodes of this extended network are also depicted in this graph, and they simultaneously represent various modules. We have investigated the enrichment of Panther biological processes using OmicsNET (refer to **Table 4**).

**Figure 6.**
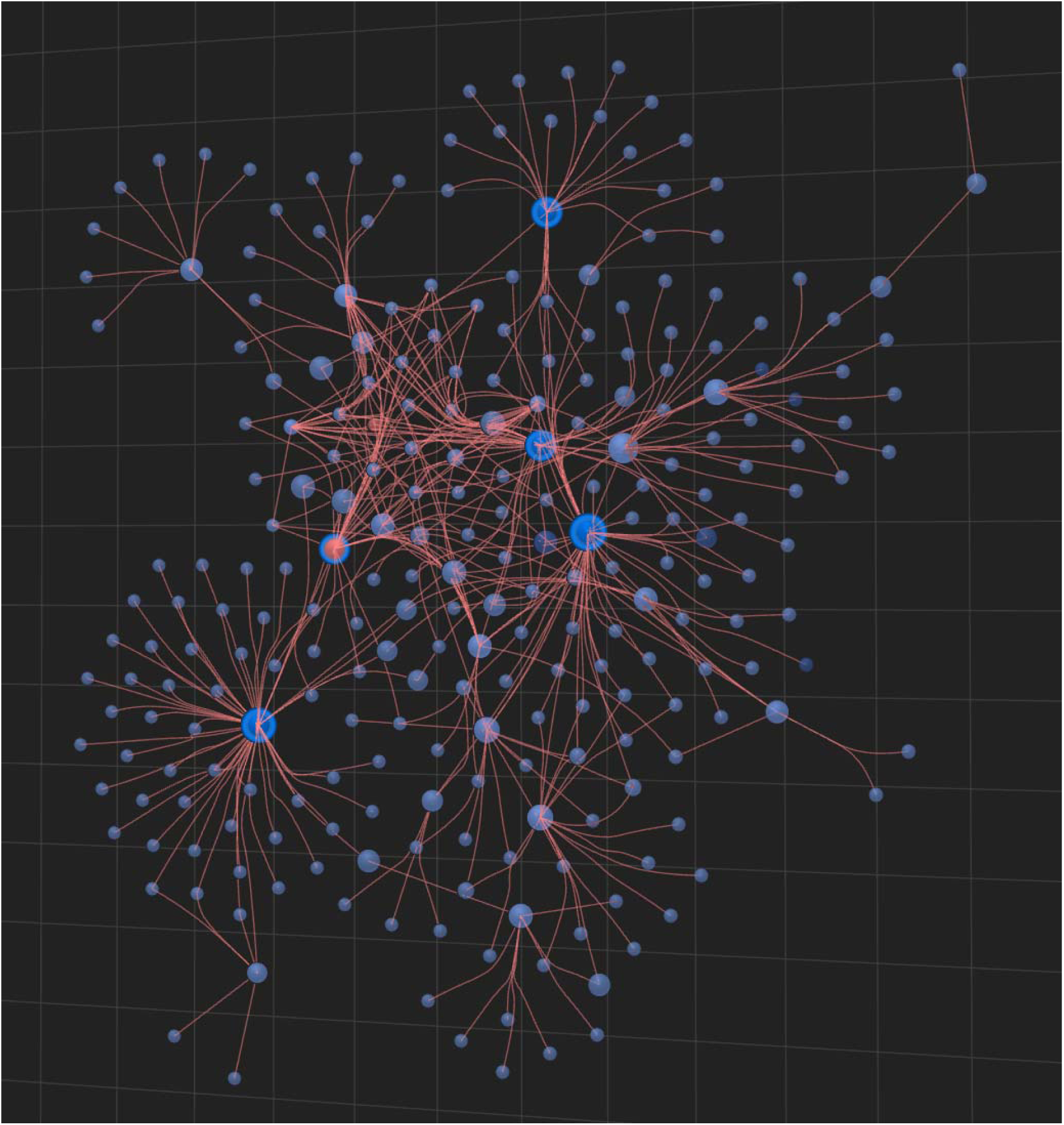
The extended network visualization of differently expressed cytokine / chemokine / growth factor / CD marker genes of major dysmood disorder. This enlarged network was constructed using OmicsNET analysis and STRING enrichment. TNF: gene ID of tumor necrosis factor-α; CCL5: chemokine ligand 5 or RANTES; CXCL12: CXC motif chemokine ligand 12 or stromal cell-derived factor 1 (SDF-1); CD4: cluster of differentiation 4; HLA-DR: Human Leukocyte Antigen – DR isotype.

**Table 4.** Panther biological processes associated with major dysmood disorder.

| **Pathway** | **Total** | **Expected** | **Hits** | **p value** | **FDR** |
| --- | --- | --- | --- | --- | --- |
| Immune response | 387 | 6.59 | 106 | 3.99E-106 | 7.74E-104 |
| Viral process | 448 | 7.63 | 48 | 2.06E-25 | 1.99E-23 |
| Immune system process | 515 | 8.77 | 44 | 2.40E-19 | 1.55E-17 |
| Cell-cell signaling | 232 | 3.95 | 30 | 2.66E-18 | 1.29E-16 |
| Antigen processing and presentation | 43 | 0.732 | 15 | 1.75E-16 | 6.81E-15 |
| Antigen processing and presentation of peptide or polysaccharide antigen via MHC class II | 16 | 0.272 | 9 | 1.04E-12 | 3.37E-11 |
| Apoptotic process | 699 | 11.9 | 40 | 8.46E-12 | 2.34E-10 |
| Negative regulation of apoptotic process | 577 | 9.82 | 32 | 3.00E-09 | 7.28E-08 |
| Cellular defense response | 59 | 1 | 11 | 3.66E-09 | 7.89E-08 |
| Cellular calcium ion homeostasis | 102 | 1.74 | 11 | 1.23E-06 | 2.39E-05 |
| Protein phosphorylation | 627 | 10.7 | 24 | 0.000169 | 0.00298 |
| Cytokine production | 31 | 0.528 | 4 | 0.00179 | 0.029 |
| Defense response to bacterium | 164 | 2.79 | 9 | 0.00197 | 0.0294 |
| Cell adhesion | 629 | 10.7 | 21 | 0.00244 | 0.0338 |

FDR: false discovery rate p correction

This table demonstrates that immune responses was the most significant process underpinning the extended network, followed by a viral process and, at a distance, antigen processing and presentation. The defense response to a bacterium was only marginally significant. The KEGG gene/protein processes that underpin the extended network were analyzed in **Table 5**. The phosphatidylinositol signaling system exhibited the most significant enrichment in the network, followed by FoxO signaling, the sphingolipid signaling pathway, and the phospholipase D signaling pathway.

**Table 5.** KEGG gene/protein processes associated with major dysmood disorder.

| Pathway | Total | Expected | Hits | P.Value | FDR |
| --- | --- | --- | --- | --- | --- |
| Phosphatidylinositol signaling system | 100 | 2.61 | 70 | 6.43E-92 | 2.16E-89 |
| FoxO signaling pathway | 294 | 7.68 | 84 | 2.05E-68 | 3.44E-66 |
| Sphingolipid signaling pathway | 189 | 4.94 | 63 | 6.63E-55 | 7.43E-53 |
| Phospholipase D signaling pathway | 102 | 2.66 | 40 | 9.53E-38 | 8.01E-36 |
| Thermogenesis | 112 | 2.93 | 40 | 8.15E-36 | 5.48E-34 |
| Chemical carcinogenesis | 201 | 5.25 | 47 | 1.38E-32 | 7.71E-31 |
| Dilated cardiomyopathy (DCM) | 38 | 0.993 | 25 | 2.47E-31 | 1.18E-29 |
| TNF signaling pathway | 107 | 2.79 | 36 | 3.71E-31 | 1.56E-29 |
| Fc epsilon RI signaling pathway | 94 | 2.46 | 33 | 2.73E-29 | 1.02E-27 |
| Viral myocarditis | 41 | 1.07 | 24 | 2.72E-28 | 9.14E-27 |
| Proximal tubule bicarbonate reclamation | 43 | 1.12 | 24 | 1.37E-27 | 4.20E-26 |
| Epstein-Barr virus infection | 170 | 4.44 | 38 | 1.53E-25 | 4.30E-24 |
| Hepatitis B | 112 | 2.93 | 32 | 3.43E-25 | 8.86E-24 |
| Transcriptional misregulation in cancer | 186 | 4.86 | 39 | 4.25E-25 | 1.02E-23 |
| JAK-STAT signaling pathway | 104 | 2.72 | 31 | 4.75E-25 | 1.06E-23 |

FDR: false discovery rate p correction

## Discussion

### Topology of the neuro-immune network in MDMD

The first major discovery of this study is that the zero and extended networks of the cytokine, chemokine, growth factor, and CD marker DEPs were primarily composed of TNF, IL-6, CXCL12, CXCL10, CCL5, CD4, CD8A, HLA-DR, and FOXP3. Consequently, chemokines and CD markers are crucial components of the immunological networks involved in MDMD. Furthermore, the findings indicate that T cell markers, such as CD4+ (T helper cells) and CD8+ (cytotoxic cells), along with the T cell activation marker HLA-DR+, are significant hotspots within the interconnected cohesive network.

The hubs of a PPI-network are of paramount importance, as they are fundamental to the network and have a significant impact on numerous nodes. Consequently, these influential nodes may be regarded as hotspots and, consequently, as legitimate drug targets. On the other hand, the information flow within the network is regulated by the bottlenecks, as they are the nodes where the network’s traffic is regulated. Consequently, both the hotspots and bottlenecks are part of the backbone of the network that, when altered, will have a modulating effect on the entire network (Maes, Plaimas et al. 2021).

Prior studies primarily examined pivotal M1 cytokines, including IL-1β, IL-6, TNF-α, and IL-8, as evidenced by several studies and meta-analyses (Maes, Meltzer et al. 1995, Ng, Tam et al. 2018, Çakici, Sutterland et al. 2020). Consequently, the findings of the current suggest that the chemokine module is at least as significant as the M1 cytokines that were introduced three decades ago (Maes, Bosmans et al. 1991). Three meta-analyses that were conducted in MDD showed that CCL2 (Köhler, Freitas et al. 2017), CCL2 and MCP-1 (Eyre, Air et al. 2016), and CCL2, CCL3, CCL11, CXCL7 and CXCL8 (Leighton, Nerurkar et al. 2018) were increased in MDD as compared with controls. Nevertheless, the results of our study show that chemokines such as CXCL12, CXCL10, and CCL5 emerge as hotspots within the network and thus as highly significant drug target candidates for the treatment of MDMD. It is important to note that the cytokine exhibiting the greatest effect size in MDMD, specifically IL-16, was neither a hub nor a bottleneck, whereas IL-6, a less significant cytokine in MDD, including in the study by (Almulla, Abbas Abo Algon et al. 2024), was a significant hub.

Furthermore, our findings indicate that growth factors or colony-stimulating factors, which are highly increased in MDMD (such as PDGF and M-CSF), do not function as a hub or bottleneck. Therefore, IL-16, CXCL27, M-CSF, and PDGF are not the primary drug targets, despite the fact that they are more significantly increased and have a larger effect size in MDMD than for example IL-6. This is all important because it indicates that prior research concentrated excessively on the effects of the M1 cytokines such, as TNF-α, IL-6, and IL-1, and did partly neglect the most important chemokines and CD makers (Maes, Meltzer et al. 1995, Köhler, Freitas et al. 2017, Ng, Tam et al. 2018, Çakici, Sutterland et al. 2020, Osimo, Pillinger et al. 2020, Islam, Sohan et al. 2023, Jayakumar, Jennings et al. 2023, Zhang, Wang et al. 2023). Consequently, the primary drug targets to treat MDMD are TNF-α, IL-6, CXCL12, CXCL10, CCL5, CD4, CD8A, HLA-DR, and FOXP3.

We have previously reviewed the mechanisms by which the activation of this cytokine/chemokine/growth factor network may induce depressive symptoms through neurotoxic effects on affective circuits in the brain, including diminished neuroplasticity and neurogenesis, axonal dysfunctions, and structural deficits in neurons (Kubera, Obuchowicz et al., 2011; Maes and Carvalho, 2018).

### Involvement of the PPI network in IRS responses

The most significant biological pathways enriched in the PPI network are an IRS response, cytokine-cytokine receptor interactions, a cellular defense response, and, in particular, a viral process that involves viral protein interaction with cytokine and cytokine receptors. This was illustrated by our different enrichment and annotation analyses. In the same vein, the network’s backbone identified hub and bottleneck nodes, which are essential components of an IRS response and a defense response against viral infections.

As a result, TNF is a most critical hotspot, and TNF signaling is a critical pathway that supports the network. During an immune response, monocytes and macrophages (as well as other immune cells) produce both TNF-α and IL-6, which are pleiotropic cytokines (Idriss and Naismith 2000, Maes, Anderson et al. 2014). TNF-α is a critical component of the host’s response to infection and is produced in response to viral infections (Winthrop, Baddley et al. 2013). Its production triggers an IRS response, which involves the recruitment of immune cells to the infectious site and the activation of macrophages (and other cells) (Kim and Solomon 2010). IL-6 is a critical factor in the acute phase response and numerous immune processes associated with MDD (Maes, Anderson et al. 2014). Additionally, the upregulation of IL-6 during infection may facilitate the survival of the virus and exacerbations (Velazquez-Salinas, Verdugo-Rodriguez et al. 2019). The chemokine network is an additional mediator of the IRS response, and chemokines are essential for the defense against viral infections (Melchjorsen, Sørensen et al. 2003). Chemokines attract white blood cells to the infectious site and stimulate them to produce pro-inflammatory cytokines (Melchjorsen, Sørensen et al. 2003). Viral infections are accompanied by elevated levels of CCL5 and CXCL10 including in the cerebrospinal fluid. CXCL12, a pre-B cell growth factor, is essential for viral infections and serves important homeostatic functions (Janssens, Struyf et al. 2018).

The activation and proliferation of CD4+ and CD8+ T cells are another prominent response to viral infection (Harty, Tvinnereim et al. 2000). CD4+ T cells facilitate the generation of virus-specific memory CD8+ T cells and maximize CD8+ T cell populations during an IRS response (Swain, McKinstry et al. 2012). CD8+ T cells maintain the antiviral response by producing chemokines and they have direct effector functions through cytotoxic T lymphocyte-mediated lysis (Swain, McKinstry et al. 2012). In MDMD, activated T cells (as indicated by increased HLA-DR+ T cell numbers and other activation markers) trigger the production of the chemokine network and consequently white blood cell recruitment to viral replication sites (Swain, McKinstry et al. 2012). Other research indicates that HLA-DR may be a valuable marker for the identification of effector T cells and the surveillance of immune responses in the context of infections (Tippalagama, Singhania et al. 2021).

The defense response to an invading pathogen comprises an IRS response that eliminates or manages the infection, and a CIRS response that mitigates collateral harm from an excessive IRS reaction (Belkaid 2008, Maes and Carvalho 2018). FOXP3+ T cells are crucial to these regulatory processes, hence averting tissue injury (Belkaid 2008). Nonetheless, when Tregs effectively maintain host homeostasis by regulating excessive immune responses, a resultant effect of this regulation is increased pathogen survival and even pathogen resistance (Scott-Browne, Shafiani et al. 2007). Moreover, Treg cells may facilitate viral replication (Belkaid 2008). Conversely, patients exhibiting diminished Treg cell counts and chronic viral infections (e.g., HCV) may manifest autoimmune processes. This is significant because MDMD is characterized by a relatively reduced level of Treg cells, which is linked to the severity of depression (Maes, Zhou et al. 2024). Furthermore, whereas Treg cells may decrease during the acute phase of severe depression, our findings indicate that in the (partially) remitted phase, the quantity of Treg cells is elevated (Maes, Nani et al. 2021). Consequently, FOXP3+ cells may play a role in viral infections or (auto)immune responses during both the acute phase and the (partially) remitted phase of MDD.

It is imperative to note that viral processes, including EBV and CMV infections, are highly significant processes that underpin the PPI network constructed in this study. It is important to emphasize that the immune-risk phenotype of bipolar depression may be influenced by human cytomegalovirus (HCMV)-specific seropositivity (Maes, Nani et al. 2021). In addition, depression due to multiple sclerosis and depression following acute coronavirus 2019 (COVID-19) disease (Long COVID) are two conditions that are distinguished by elevated IgG/IgA/IgM levels against deoxyuridine 5′-triphosphate nucleotidohydrolase (dUTPAses) of EBV and HHV-6 (Maes, Almulla et al. 2024, Maes, Almulla et al. 2024). The latter findings indicate the presence of HHV-6 and/or EBV replication and reactivation in association with depressive symptoms.

### Other relevant pathways

Other pathways that are enriched in the PPI network constructed around the DEPs of MDMD are the NF-κB pathway, the TLR pathway, the phosphatidylinositol signaling system, the sphingolipid signaling pathway, the phospholipase D signaling pathway, and the FOXO signaling pathway. MCODE analysis discovered one molecular complex comprising activation of the cytokine network and viral protein interactions combined with regulation by RELA.

Previously, we used annotation and enrichment analyses on a DEP (cytokine) network in severe MDD (Maes, Rachayon et al. 2022). Apart from increased TNF signaling, hyper-responsive NF-κB and TLR pathways were major molecular pathway underpinning MDMD (Maes, Rachayon et al. 2022). Recent papers show that NF-κβ RNA and TLR-4 RNA and protein levels are significantly increased in peripheral blood mononuclear cells of patients with MDD (Kéri, Szabó et al. 2014). The prefrontal cortex of individuals with MDD shows increased mRNA expression of TLR-4 and TLR-3 (Pandey, Rizavi et al. 2014). A recent review demonstrated an increased expression of TRIF (TIR domain-containing adaptor molecule 1), MYD88 (an innate immune signal transduction adaptor), TLR-3/4, and NF-κB in individuals diagnosed with MDD (Mariani, Cattane et al. 2021). TRIF and MYD88 are both key elements leading to NF-κB pathway activation. Similar findings were reported by Elovainio et al. (2015) who discovered that individuals with severe depression exhibit gene-set pathways linked to the TLR pathway and the NF-κβ pathway (Elovainio, Taipale et al. 2015).

Upon the engagement of a ligand to certain TLR complexes, NF-κB can be transported to the nucleus, resulting in the activation of transcription for pro-inflammatory cytokines and other actors leading to a primed inflammasome. Viral pathogen-associated molecular patterns (PAMPs), which include viral proteins and nucleic acids, are sensed by toll-like receptors (TLRs), including TLR2, TLR3, TLR4, TLR7, TLR8, and TLR9 (Lester and Li 2014). Consequently, the stimulation of these TLRs initiates distinct signaling pathways, which ultimately result in an antiviral response that is orchestrated by the increased production of interferons (IFNs), pro-inflammatory cytokines and chemokines (Lester and Li 2014). These antiviral responses are influenced by two significant signal transduction pathways: MYD88 and TRIF, which both result in the activation of NF-κB. It is imperative to emphasize that the NF-κB complex (which includes RELA and REL) was the most significant transcription factor associated with our PPI network, as demonstrated by our TRRUST transcription factor enrichment analysis. The NF-κB pathway induces proinflammatory cytokines and contributes to an increase in IFN production (Lester and Li 2014). The NF-κB subunit RELA exhibits a dual function in viral infections. Firstly, RELA traverses the nucleus to maintain baseline and autocrine IFN-β signaling in uninfected cells, a process crucial to immune responses against viral infections. Secondly, RELA amplifies inflammatory and adaptive immune responses following infection and may facilitate the survival of infected cells throughout this phase (Basagoudanavar, Thapa et al., 2011).

Cell signaling, cytoskeleton regulation, organelle identity, and membrane dynamics are all significantly influenced by phosphatidylinositol phosphate kinases. Numerous viral infections target this pathway (Beziau, Brand et al. 2020). Sphingolipids, which are crucial components of cell membranes, are involved in cell signaling, and their metabolites are essential for the host’s defense against viral infections (Thomas, Samuel et al. 2023). The IRS response to viral infections is significantly influenced by perturbations in sphingolipid metabolism (Janneh, Kassir et al. 2021). Ceramides, a type of sphingolipid, may also function as a receptor for viral entry into host cells, thereby facilitating viral replication (Thomas, Samuel et al. 2023). Also, phospholipase D may facilitate the entry of certain viruses into host cells and contribute to viral infections (Oguin, Sharma et al. 2014). The FOXO signaling pathway is responsible for the regulation of IRS responses, oxidative stress, Th1 activation, and the enhancement of FOXP3 and Treg cell formation and, consequently, the maintenance of cellular quality (e.g. redox and inflammatory homeostasis) (Cabrera-Ortega, Feinberg et al. 2017). The FOXO regulatory pathway is crucial in viral infections, including influenza, as it mediates anti-inflammatory and anti-apoptotic mechanisms (Wu, Zhang et al. 2019).

## Conclusions

The backbone nodes and, consequently, the primary drug targets for MDMD, as determined by the PPI network analysis conducted in the present investigation, are TNF, IL6, CXCL12, CXCL10, CCL5, CD4, CD8A, HLA-DR, and FOXP3. A cellular or physiological defense response, an IRS response, cytokine-cytokine receptor interactions, and, in particular, a viral process that involves viral protein interaction with cytokine and cytokine receptors are the most significant biological pathways or processes enriched in the PPI network. MCODE analysis disclosed a significant molecular component composed of immune and viral processes and “regulated by RELA”. It seems that viral mechanisms could play a fundamental role in the activation of T cells, as well as in activation of the cytokine and chemokine networks linked to MDMD.

The findings of the present study indicate that future research on the immunological pathogenesis of MDMD (and MDD) should concentrate on the comparatively underexamined immune indicators within the chemokine network, cell surface CD or intracellular markers, evaluated using flow cytometry, and the NF-κB pathway including its subunit RELA. Future research should investigate whether and which viral infections are linked to the onset of MDD/MDMD, or whether viral reactivation is associated with the recurrence of illness. Three candidates that necessitate examination are HCMV, EBV, and HHV-6. In the interim, research should investigate which medications can be employed to normalize the immune system aberrations in MDMD, which include the chemokine network (particularly CCL, CXCL10, and CXCL12), T cell activation (CD4, CB8, and HLA-DR), and attenuated FOXP3 expression.

## Data Availability

MM and AFA designed the study. MM, KP and AS conducted the studys statistical analysis. MM. wrote the first draft of the paper AFA, KP, AS, YZ, and JL further edited the paper. All authors have read and approved the final manuscript. AS was funded by National Science, Research and Innovation Fund (NSRF), and King Mongkuts University of Technology North Bangkok (Project no. KMUTNB-FF-67-B-24).

## Acknowledgments

The authors express gratitude to the personnel of Al-Hakeem General Hospital, Najaf, Iraq and Chulalongkorn Hospital in Bangkok, Thailand for their contributions to data collecting.

## Ethical approval and consent to participate

This study involves a secondary data analysis using open, deidentified, and non-coded data sets, so exempting it from IRB approval as non-human subjects’ research. The utilized data are derived from two case-control investigations (Almulla, Abbas Abo Algon et al. 2024, Maes, Zhou et al. 2024). Both case-control studies received approval from the local Institutional Review Boards, and all participants furnished signed informed consent before their participation in the study. The Institutional Review Board of the College of Medical Technology of the Islamic University of Najaf, Iraq, approved the study under Document No. 18/2021. The research procedure by Maes et al. (2024) was sanctioned by the Institutional Review Board of Chulalongkorn University, Faculty of Medicine, Bangkok, Thailand (#528/63).

## Author’s contributions

MM and AFA designed the study. MM, KP and AS conducted the study’s statistical analysis. MM. wrote the first draft of the paper AFA, KP, AS, YZ, and JL further edited the paper. All authors have read and approved the final manuscript. AS was funded by National Science, Research and Innovation Fund (NSRF), and King Mongkut’s University of Technology North Bangkok (Project no. KMUTNB-FF-67-B-24).

## Availability of data

All data used in the study are made available in the Tables and Figures.

